# Validation of a Non-invasive Prenatal Test for Fetal RhD, C, c, E, Kell and FyA Antigens

**DOI:** 10.1101/2023.03.16.23287355

**Authors:** Brian Alford, Brian P. Landry, Sarah Hou, Xavier Bower, Anna M. Bueno, Drake Chen, Brooke Husic, David E. Cantonwine, Thomas F. McElrath, Jaqueline Carozza, Julia Wynn, Jennifer Hoskovec, Kathryn J. Gray

## Abstract

We developed and validated a next generation sequencing (NGS) based NIPT assay using quantitative counting template (QCT) technology to detect RhD, C, c, E, Kell, and Fy^a^ fetal antigen genotypes in the diverse U.S. population. The assay quantifies paternally derived fetal antigen cell-free DNA molecules after calibration to fetal fraction and a reference gene. The assay correctly determined fetal antigen status for 1061 preclinical samples with an analytical sensitivity of 100% (95% CI: 99-100%) and analytical specificity of 100% (95% CI: 99-100%). The assay showed a clear separation between antigen detected and not detected for 15,939 clinical plasma samples in a general population setting, with an estimated clinical sensitivity of 99.6%-100%. The precision of the assay in which two replicate plasma samples were independently analyzed was 99.9% for 1683 clinical samples. Moreover, a fetal antigen determination could be made for samples with *RHDΨ*, a variant more common among RhD-negative Black individuals. The NIPT results were 100% concordant with neonatal antigen genotype/serology for 23 RhD negative pregnant individuals and 12 other antigen evaluations in 4 alloimmunized pregnant individuals. This NGS-based fetal antigen NIPT assay had excellent performance in a validation study of samples from a diverse U.S. population for fetal fractions as low as 1.1% and as early as 10 weeks of gestation, without the need for a sample from the biological partner. Implementation of NIPT for the detection of fetal antigen in RhD-negative and alloimmunized pregnant individuals will streamline care and reduce unnecessary treatment, monitoring and patient anxiety.

## INTRODUCTION

Hemolytic disease of the fetus and newborn (HDFN) is a serious blood disorder caused by the destruction of a fetus or neonate’s red blood cells (RBCs) by the immunoglobulin G (IgG) antibodies of the pregnant person. HDFN occurs when an antigen is expressed on the fetal RBCs but not on the RBCs of the pregnant person and the fetal RBCs gain access to the pregnant person’s circulation and induce an immune response. HDFN can cause severe fetal anemia which in turn may lead to fetal hydrops, heart failure, hyperbilirubinaemia and associated kernicterus (toxic levels of bilirubin in the brain), and perinatal death.

Historically, the most common cause of HDFN was Rhesus factor (Rh) incompatibility, i.e., when the pregnant person is RhD-negative and the fetus is RhD-positive. However, the introduction of anti-RhD immunoglobulin in the 1970s drastically reduced the risks associated with Rh incompatibility. Sensitization (a pregnant person’s immune response to RhD-positive RBCs) and RhD-associated HDFN has decreased from 16% of RhD-negative pregnant individuals to less than 1% of RhD-negative pregnant individuals [1]. The current clinical standard of care in the U.S. is to administer prophylactic anti-RhD immunoglobulin to all RhD-negative pregnant people to prevent sensitization; however, this intervention is unnecessary in approximately 40% of pregnant individuals, as they carry an RhD-negative fetus. [2]

In addition to RhD, other antigens are associated with HDFN, with approximately 0.5-1% of pregnancies now complicated by alloimmunization[1]. The risk of alloimmunization is greater among populations that receive regular blood transfusions, such as individuals affected with sickle cell disease [3–5]. In the U.S., the standard of care is antibody screening in the first trimester of pregnancy and if antibodies associated with HDFN are identified, additional testing is indicated to determine the HDFN risk for the fetus [1]. This includes serological or genetic evaluation of the partner, when possible, and amniocentesis to determine fetal antigen genotype. Determination of fetal antigen genotype reduces unnecessary monitoring and treatment of pregnancies with concordant antigen status that are not at risk for HDFN. However, barriers to partner antigen screening and reluctance to undergo an invasive procedure leads to lack of knowledge about the fetal antigen status in many pregnancies which then must be treated as high-risk for HDFN. In approximately 35-50% of pregnancies with alloimmunization, the fetus is antigen positive and at risk for HDFN, however, in the remaining pregnancies, the fetus is negative for the corresponding antigen and therefore is not at risk [5].

Non-invasive prenatal testing (NIPT) of cfDNA can be used to determine fetal antigen genotype for the prediction of fetal antigen phenotype. Many European countries have adopted this practice to determine administration of anti-RhD immunoglobulin and to guide pregnancy management of alloimmunized pregnant individuals [2, 6–10]. However, European-based assays are logistically challenging for U.S.-based patients, often require testing to be delayed until after 20 weeks’ gestation, and have high rates of inconclusive results, particularly for people of non-European ancestry [1, 2, 11–13].

A challenge of the NIPT assays used in Europe is that they employ qualitative polymerase chain reaction (PCR) technology under the assumption that an RhD-negative fetus will be homozygous for the *RHD* gene deletion, which is the most common genotype that results in an RhD-negative phenotype [14]. However, up to 30% of Black RhD-negative individuals have non-*RHD* gene deletion variants, such as *RHDΨ*, that cannot be detected or are incorrectly identified as fetal RhD-positive by these assays [15–17]. Additionally, qualitative NIPT assays do not directly measure the fraction of cfDNA of fetal origin, but rather rely on the detection of Y chromosome genes or a single autosomal reference gene to confirm the presence or absence of fetal cfDNA, thereby resulting in a fetal sex bias inaccuracy and an increased potential for false negatives especially if tested too early in pregnancy when the fraction of cfDNA of fetal origin is lower [18–20].

The American College of Obstetricians and Gynecologists (ACOG) noted that NIPT could be an effective and attractive strategy for the management of RhD-negative and alloimmunized pregnancies if the inclusivity, feasibility, and cost-effectiveness of cfDNA tests are addressed for the U.S. population [1]. The application of the recently developed quantitative counting template (QCT) technology to NIPT addresses the barriers of inclusivity and feasibility for the U.S. population. The technology is capable of both accurately quantifying genes of interest as well as sequencing variants using Next Generation Sequencing (NGS). QCT technology has previously been validated for NIPT of autosomal recessive conditions in cfDNA [21, 22].

Here we describe the application of QCT technology NIPT for the detection of the fetal genotypes that encode for the RhD, C, c, E, Kell, and Duffy (Fy^a^); RBC antigens associated with HDFN [1], and show that this assay demonstrates excellent performance even at fetal fractions as low as 1.1% and as early as 10 weeks of gestation, with a sensitivity and specificity of 99.6%-100% across multiple sets of validation. This performance is achieved by the combination of QCT technology with a genome-wide assessment of polymorphic locations that determine cfDNA fetal molecule count [22]. Moreover, the quantitative sequencing-based design of the fetal antigen NIPT described herein enables the detection of fetal antigen status due to the common genotypes as well as non-deletion *RHD* gene variants, thus improving clinical utility for the diverse U.S. population and avoiding the necessity of a partner sample.

## Methods

### Assay overview and design

To detect the genotype of fetal antigens RhD, C, c, E, Kell, and Fy^a^ in pregnant individuals, a highly-multiplexed PCR based NIPT assay with a NGS readout that employs QCT technology was developed. In the U.S., approximately 26% of RhD-negative Black individuals have one or more *RHDΨ* variants, and up to 15% have an *RHD-CE-D* hybrid gene [15, 16]. Therefore, it is imperative that an NIPT assay used in the U.S. population can predict fetal Rh phenotype when these variants are present. In order to enable the detection of the fetal RhD genotype (and therefore the expected phenotype) in the presence of the common *RHD* gene deletion, as well as variants associated with an RhD-negative phenotype such as *RHDΨ* and the *RHD-CE-D* hybrid gene, the assay is comprised of five amplicons across the RHD gene that amplifies regions that are unique between the wild type RHD gene, the RHCE homolog gene (exons 7 and 10) and the *RHDΨ* RHD gene variant (exons 4 and 5) (Figure 1a) [16].

**Figure 1.**
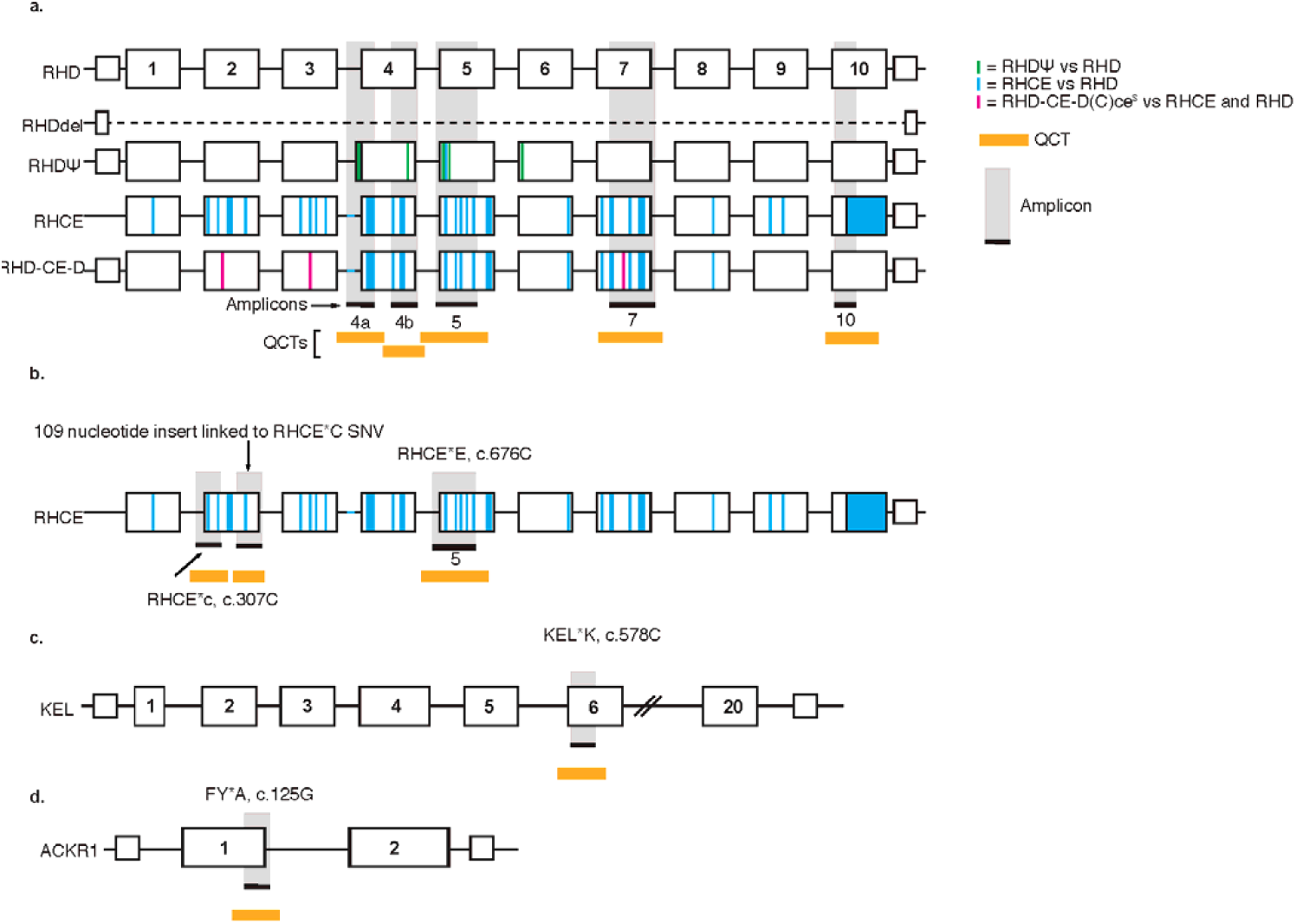
Schematic of the genes associated with the antigens, amplicons, and quantitative counting templates (QCTs). QCTs quantify the molecular count for each genomic loci. **A)** The wild type *RHD* gene is shown along with its common variants, *RHD* gene deletion and *RhDΨ* variant, and the homologous *RHCE* gene and *RHD-CE-D* hybrid gene. Differences between these variants and homologs are highlighted as indicated. **B)** Schematic of the *RHCE* gene, single nucleotide variants (SNVs) associated with RHCE*C (c.307C), RHCE*c (c.307T) and RHCE*E (c.676C) genotypes. The amplicon for the RHCE*C quantifies a 109 nucleotide insert highly linked to RHCE*C. The RHCE*E (c.676C) is located in the in amplicon 5 of the RhD assay **C)** Schematic of the *KEL* gene and KEL*K (c.578C) SNV **D)** Schematic of the *ACKR1* gene and FY*A (c.125G) SNV **E)** equations for absolute expected molecules (AEM) and calibrated fetal antigen fraction (CFAF)

The remaining RBC antigens (C, c, E, Kell, and Fy^a^) result from single nucleotide variants (SNVs) *RHCE*C, RHCE*c, RHCE*E, KEL*K* and *FY*A*, respectively. A single amplicon was designed for each SNV based on previously published antigen genotyping assays for antigen phenotype prediction. Specifically,

- the amplicon for C antigen phenotype quantifies a 109 nucleotide insert linked to SNV *RHCE*C* [23] (**Figure 1b**),
- the amplicon for the c antigen phenotype directly amplifies SNV *RHCE*c* [9] (**Figure 1b**),
- the *RHCE*E* SNV [24], which corresponds to the e antigen phenotype, is located in amplicon 5 of the RhD antigen assay (**Figure 1b**), and
- the Kell and Fy^a^ amplicons target the SNVs that determine expression of these two antigens (*KEL*K* and *FY*A*, respectively) [24] (**Figure 1c, 1d**).

A quality control (QC) threshold of the absolute expected number of fetal molecules (AEM) from the locus of interest based on the cfDNA fetal fraction in a given sample was set for each antigen genotype to maximize sensitivity. To ensure that the assay sensitivity for an individual sample does not fall below 98%, even for the samples at the lowest threshold of fetal molecules, the AEM QC thresholds for each antigen genotype were determined by modeling fetal molecules with a Poisson distribution and were calibrated and verified through analytical validation.

To ensure the specificity of the assay, a calibrated fetal antigen fraction (CFAF) was measured for each antigen. The CFAF is equal to the observed fetal molecules for the antigen of interest divided by the AEM. In an antigen-negative pregnant person, any cfDNA from that antigen will be paternally derived; therefore, fetuses that are antigen-positive and antigen-negative would have expected CFAFs of 100% and 0%, respectively. CFAF is therefore corrected to normalize values around 100% for antigen-positive fetuses based on amplification efficiencies for individual antigen loci. For the RhD antigen, a CFAF is computed for each amplicon and the second highest measurement is used as a summary CFAF to reduce the risk of false negatives that can be caused by any potential amplification outliers. Such outliers can occur when rare SNVs coincide with the primer regions.

An intermediate no-call CFAF range was determined for each antigen genotype to prevent any false positives due to spurious issues such as sequencing noise, amplification errors, or contamination. The CFAF call thresholds were determined based on the CFAF distributions of 8,992 clinical plasma samples from pregnant individuals not used elsewhere in validation (**Table S1**).

### Preparation of preclinical samples

A PCR-based quantitative NIPT assay was designed for the detection of RhD, Kell, C, c, E, and Fy^a^ antigens. Preclinical samples were created from twelve parent-child genomic DNA pairs with known antigen genotypes purchased from Coriell. At least one parent antigen-negative/child antigen-positive pair and one parent antigen-negative/child antigen-negative pair were created for each antigen to mimic the genotype combinations of clinical samples. To make the preclinical samples, DNA was sheared to an average size of 175bp using a Covaris S220 focused-ultrasonicator to mimic cfDNA fragmentation. Sheared DNA was diluted to a concentration of 0.236 ng/μL or 0.471 ng/μL and the parent-child DNA was mixed to mimic fetal fractions ranging from 1.5% to 12%. Samples were replicated to create at least 93 samples for each antigen with an overweighting of lower fetal fractions to assure assay performance at low fetal fractions (**Table S2**).

### Selection and preparation of plasma samples from pregnant individuals with unknown fetal antigen genotypes and phenotypes

Three independent batches of clinical plasma samples from pregnant individuals with unknown fetal antigen genotype or phenotype were used in the development and initial validation of the NIPT assay. The batch of 8,992 clinical plasma samples was used to calibrate the assay as discussed in the assay overview section. A second batch of 15,939 plasma samples was used to validate the assay after initial validation on preclinical samples. A third batch was used to evaluate assay precision in which two plasma samples were analyzed independently from 1,683 patients.

Plasma samples from all pregnant individuals who had a negative genotype for at least one RBC antigen were included in these analyses, which would be the genotype of an individual alloimmunized for that antigen. Samples were anonymized and included in the study regardless of the pregnant individual’s alloimmunity status, which was unknown.

The plasma samples were processed according to the clinical fetal antigen NIPT assay pipeline to calculate the CFAF used to predict fetal antigen status. First, cfDNA was purified from plasma and size separated from genomic DNA using magnetic bead-based purification. Sequencing libraries were prepared by conducting one round of multiplexed PCR and one round of PCR-based indexing on 10 μL of purified DNA. Libraries were sequenced on an Illumina NextSeq 2000 sequencer with 1×101 sequencing. The fetal fraction of the cfDNA was determined by quantifying up to nine different polymorphic locations from a total of 64 locations throughout the genome and CFAF were computed and the fetal genotype calls were made.

For the first batch of plasma samples, CFAF distributions were used to calibrate the assay and determine the appropriate QC and no-call thresholds. For the second batch, the NIPT fetal antigen status prediction was made for all antigens where the pregnant individuals was antigen negative and as fetal/neonatal antigen genotype or phenotype were unknown for these pregnancies, a model was fit to estimate the sensitivity of the assay based on the separation of positive and negative calls (**Supplementary Methods**). For the third batch, the independent calls were compared across two separate plasma samples to determine the precision of the assay.

### Selection and preparation of plasma samples from pregnant individuals with known fetal antigen

Clinical samples from pregnant individuals with known fetal antigen genotype and/or phenotype were obtained from two sources: (1) the LIFECODES biobank at Brigham and Women’s Hospital (Boston, MA) and (2) the UNITY fetal antigen patient registry study at BillionToOne. LIFECODES, an IRB-approved biorepository, includes banked plasma samples from pregnant individuals (mean gestational at entry of 11.2 weeks). Plasma samples were selected from the LIFECODES that had been banked at -80C° for less than 2 years old; from serological RhD-negative pregnant individuals; and had known antigen serology in the resulting neonate without known fetal or neonatal abnormalities.

Twenty-five RhD-negative pregnant person plasma samples were obtained. Neonatal serology was blinded to the individuals performing the NIPT assay until the NIPT results for the samples were available. Libraries were sequenced on an Illumina MiSeq sequencer with 1×150 sequencing. The cfDNA fetal fraction was determined in the same manner described in the previous methods section.

Two of the 25 samples were excluded because they did not pass QC metrics: one due to the low fetal fraction and the other due to the patient having a history of weak D phenotype (conflicting RhD-negative and RhD-positive clinical serology results). The remaining 23 samples were analyzed by the NIPT assay, the neonatal serology was unblinded, and then assessed for concordance of NIPT fetal Rh status with neonatal Rh serology.

The UNITY fetal antigen patient registry study is an IRB-approved study eligible for pregnant individuals who have antigen NIPT assay as part of their clinical care. The study protocol involves the collection of cord blood samples or buccal swabs from the neonate that results from the pregnancy. The samples are de-identified and sent to Grifols laboratory for antigen genotyping on the FDA-approved ID CORE EX platform which genotypes the sample for C, c, E, Kell, and Fy^a^ using the same SNVs as the NIPT assay [25]. Pregnant person and neonatal medical records including antibody titers and neonatal Rh serology are collected to confirm pregnant person alloimmunization status and neonatal Rh phenotype. A sample from a pregnant individual alloimmunized for one or more antigen included on the NIPT assay was selected from the UNITY study for the current analysis (n=4). The fetal antigen NIPT results were compared to the Grifols-reported antigen genotype and predictive phenotype to determine concordance.

### Statistical analysis

Sensitivity and specificity were computed for the analytical samples and biobank samples. Clopper-Pearson 95% confidence intervals are reported. Please see Supplementary Methods for more details.

## Results

### Validation of the NIPT assay on preclinical samples

The initial validation of the NIPT assay was completed on samples with established ground truth, preclinical samples made of genomic DNA from “parent” and “child” sources (see Preparation of preclinical samples) (**Table S2**). NIPT for the detection of fetal RhD antigen genotype was first assessed with four parent-child DNA pairs mixed to mimic RhD-negative/RhD-positive and RhD-negative/RhD-negative pregnant person/fetus statuses at a 12% fetal fraction. The fetal antigen NIPT assay demonstrated 100% accuracy on these samples for the identification of the fetal RhD antigen genotype.

To establish performance of the assay across different fetal antigens and a range of fetal fractions, an additional twelve preclinical samples were made for the six antigens at fetal fractions ranging from 1.5 to 12%. These samples were weighted toward lower fetal fractions to challenge the performance of the assay and demonstrate that the assay would make accurate calls even at early gestational age. The NIPT assay correctly determined fetal antigen status with an overall analytical sensitivity 100% (95% CI: 99-100%) and specificity of 100% (95% CI: 99-100%), with a no-call rate of 1.5%, across 1061 samples with fetal fraction as low as 1.5% (**Figure 2, Table 1**).

**Figure 2.**
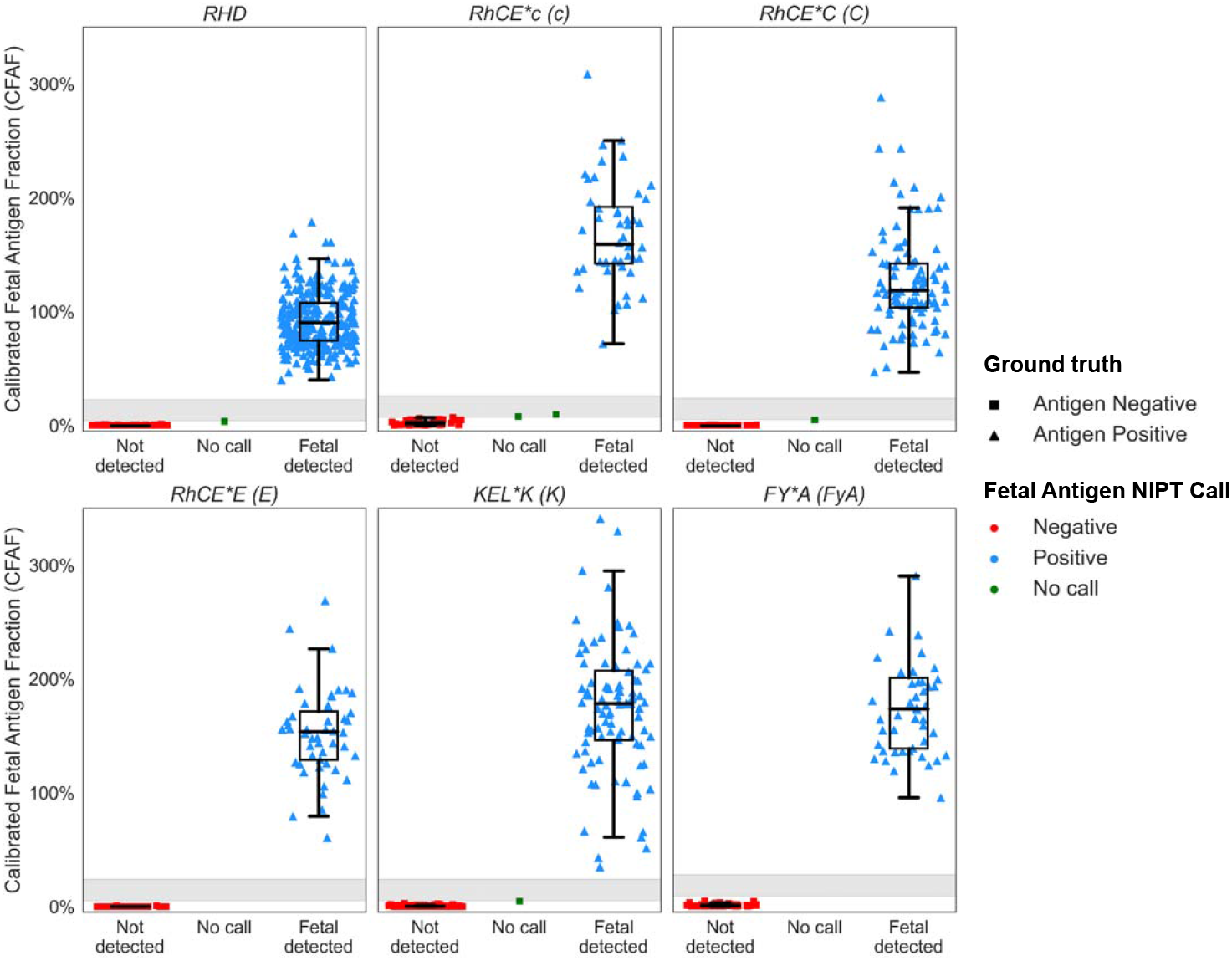
Calibrated fetal antigen fraction (CFAF)s and predicted fetal antigen status for preclinical samples. Samples were made from sheared genomic DNA samples from replicates of parent-child pairs with an antigen negative parent/ antigen positive fetus (triangle) and antigen positive parent/ antigen negative child pairs (squares) for each antigen type. Parent-child samples were mixed to mimic fetal fractions ranging from 1.5-12%. Samples with antigen detected are blue, samples with antigen not detected are red and no-call samples (CFAF intermediate range) are green. Row 1: RhD, RHCE*C and RHCE*c, Row 2: RHCE*E, KEL*K and FY*A.

**Table 1.** Sensitivity, specificity of the NIPT assays on pre-clinical samples made from genomic DNA sheared and mixed to mimic cfDNA (Table S2).

| Antigen | Positive N | Positive<br>No Call | Sensitivity | 95% CI | Negative N | Negative<br>No Call | Specificity | 95% CI |
| --- | --- | --- | --- | --- | --- | --- | --- | --- |
| RhD | 264 | 0 | 100% | (99-100%) | 191 | 1 | 100% | (98-100%) |
| RhCE*C (C) | 96 | 0 | 100% | (96-100%) | 43 | 5 | 100% | (92-100%) |
| RHCE*c (c) | 48 | 0 | 100% | (93-100%) | 46 | 2 | 100% | (92-100%) |
| RHCE*E (E) | 48 | 0 | 100% | (92-100%) | 38 | 7 | 100% | (94-100%) |
| KEL*K (K) | 96 | 0 | 100% | (96-100%) | 95 | 1 | 100% | (96-100%) |
| FY*A (FyA) | 48 | 0 | 100% | (93-100%) | 48 | 0 | 100% | (93-100%) |
| Total Fetal Antigen | 600 | 0 | 100% | (99-100%) | 461 | 16 | 100% | (99-100%) |

Clopper-Pearson 95% confidence intervals (95% CI), No Call samples had a calibrated fetal antigen fraction (CFAF) in the intermediate zone

### Evaluation of the NIPT assay on clinical samples with unknown fetal antigen genotypes and phenotypes

Next, to demonstrate the ability of the assay to assess fetal antigen genotype on clinical plasma samples, NIPT was performed on plasma obtained from 15,939 pregnant individuals with an average gestational age of 14.1 weeks (range 10.0-37.9) and a fetal fraction of 8.9% (range 1.6-37.8%) (**Table S3**) for which alloimmunization status and fetal antigen genotype/phenotype was unknown (see Selection and preparation of clinical samples with unknown fetal antigen genotypes and phenotypes). Plasma samples from the clinical patients were analyzed for fetal antigen genotype status for all antigens for which the pregnant person was genotype-negative. This sample selection is consistent with the clinical application of the assay where alloimmunized pregnant persons are genotype-negative for the antigen(s) for which they are alloimmunized. This resulted in a total of 40,596 NIPT assays on the 15,939 patients for the six antigens. The QC failure due to low numbers of measured fetal molecules, i.e., AEM, was less than or equal to 3.0% for all antigens (**Table S4**).

Of the NIPT-analyzed samples, 99.5% (N=40,406) had an informative fetal antigen genotype result for one or more fetal antigens according to the previously established intermediate, no-call range. The proportion of samples in the CFAF in the no-call range was less than 1% for any given antigen (**Figure 3a, S1a, Table S4**). The CFAF values were independent of fetal fraction (**Figure S1b**). As the fetal/neonatal antigen genotype was unknown for these samples, the samples with a predicted fetal antigen-positive genotype by NIPT were modeled to estimate the clinical sensitivity of the assay. The modeled estimated sensitivity was greater than 99.5% for all antigens (**Table S5, Figure S2**). The overall no call rate (both quality control fail and intermediate range) across all antigen assays was 2.4%.

**Figure 3ab.**
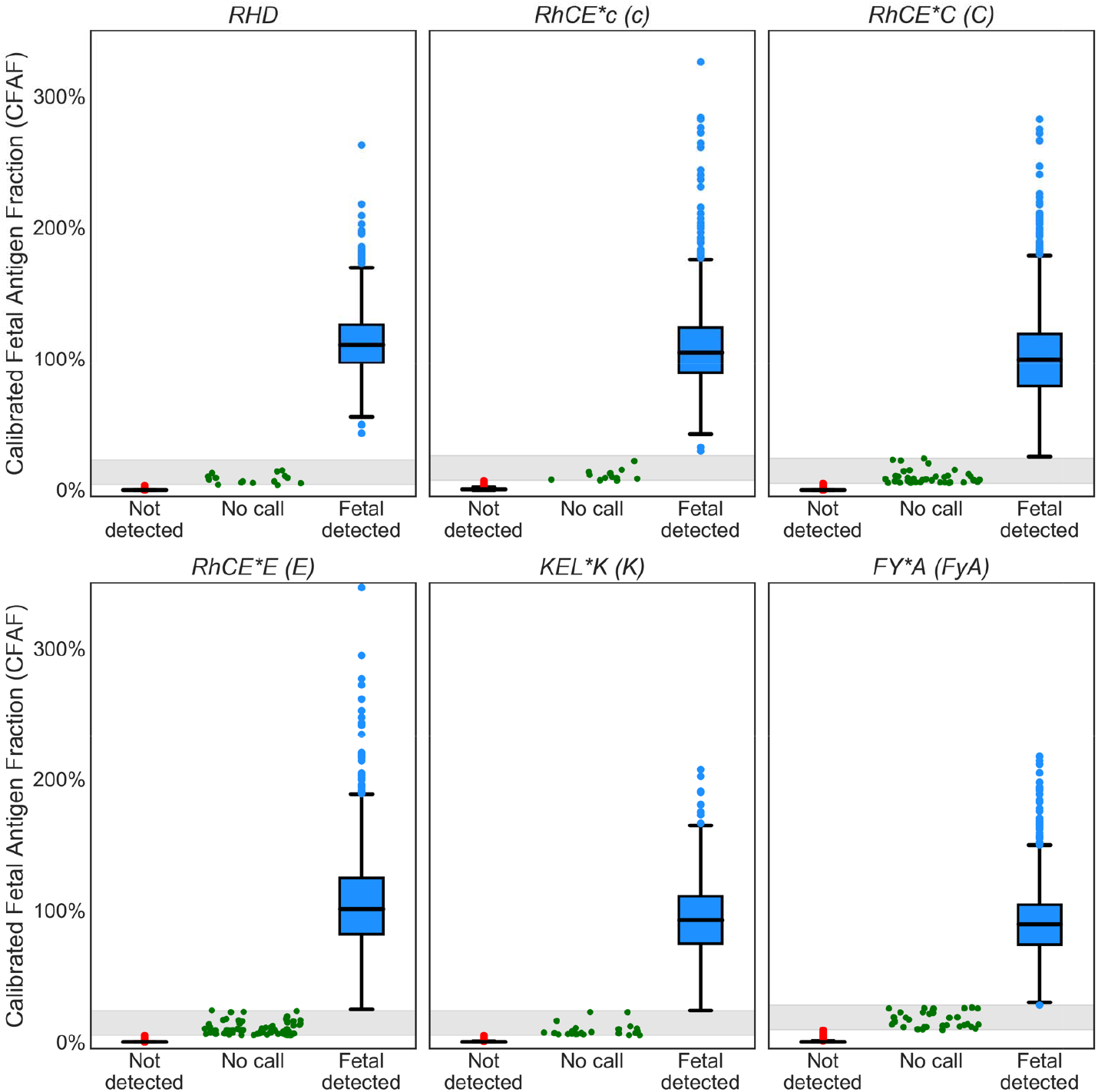
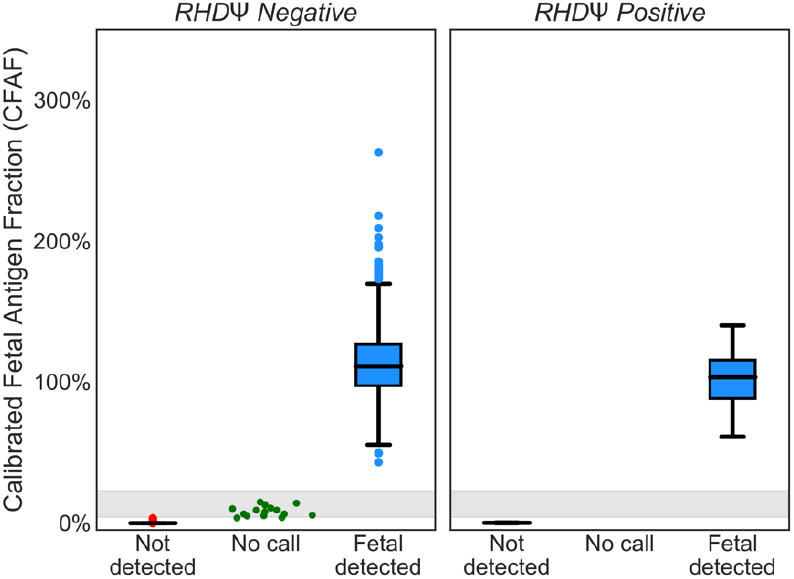
Calibrated fetal antigen fraction (CFAF)s and predicted fetal antigen status for clinical samples. Pregnant person antigen status was determined by NGS of cfDNA and fetal antigen status was reported for all samples where the pregnant person was identified to be negative for the antigen to mimic the clinical use of this assay. Samples are not unique across plots; the same sample was used for multiple assays if the pregnant person was antigen negative for multiple antigens. Antigen detected is indicated in blue, antigen not detected is indicated in red and no-call is indicated in green. **a**. Row 1: RhD, RHCE*C and RHCE*c, Row 2: RHCE*E, KEL*K and FY*A. **b**. The fetal RhD prediction for the 510 without clinical samples where the RhDΨ variant was not detected (panel 1) and the clinical samples where RhDΨ variant was detected (panel 2). In qualitative RhD NIPT assays, samples with RhDΨ variant result in a no-call or inconclusive result.

There were 56 samples (3.6% of RhD-negative samples) for which the *RHDΨ* variant was present (i.e., in pregnant person and/or fetal cfDNA). In 15, the fetus was predicted to have an RhD-negative phenotype and in 41 the fetus was predicted to have an RhD-positive phenotype (**Figure 3b, S1c**). These samples with the *RHDΨ* variant would have resulted in a no-call or in correct fetal Rh phenotype prediction for other, qualitative NIPT assays because of the inability to quantify the *RHDΨ* cfDNA and therefore predict fetal phenotype [14, 26].

Precision is another important metric that correlates with test accuracy. To evaluate analytical precision, the concordance of NIPT results for a unique set of 1,683 retained plasma samples with two remaining samples (3,366 samples) was assessed. As with the analysis on other clinical plasma samples, NIPT was completed for all antigens in which the pregnant individual was genotype-negative, resulting in a total of 3,921 NIPT analyses. All but 5 NIPT results were concordant across the two NIPT assays, resulting in a precision of 99.9% (95% CI: 99.7%-100%) (**Table S6**).

### Clinical validation on biobank samples with known serology results

Finally, NIPT was completed on 27 plasma samples where the fetal/neonatal antigen phenotype and or genotype was known to enable evaluation of the accuracy of the assay on plasma samples. The 23 blood samples from the LIFECODES biobank and four samples from the UNITY fetal antigen patient registry study had fetal fractions ranging from 1.1% to 37.8% (**Table S7**). The NIPT-predicted fetal antigen genotype was consistent with the known neonatal/fetal antigen genotype or phenotype for all samples. The concordance evaluation included twelve fetuses correctly identified as RhD-positive and eleven fetuses correctly identified as RhD-negative, resulting in a clinical sensitivity of 100% (95% CI: 73.5%-100.0%) and a clinical specificity of 100% (95% CI: 71.5%-100.0%) for the detection of fetal RhD status (**Figure 4a, Table S8ab**).

**Figure 4ab:**
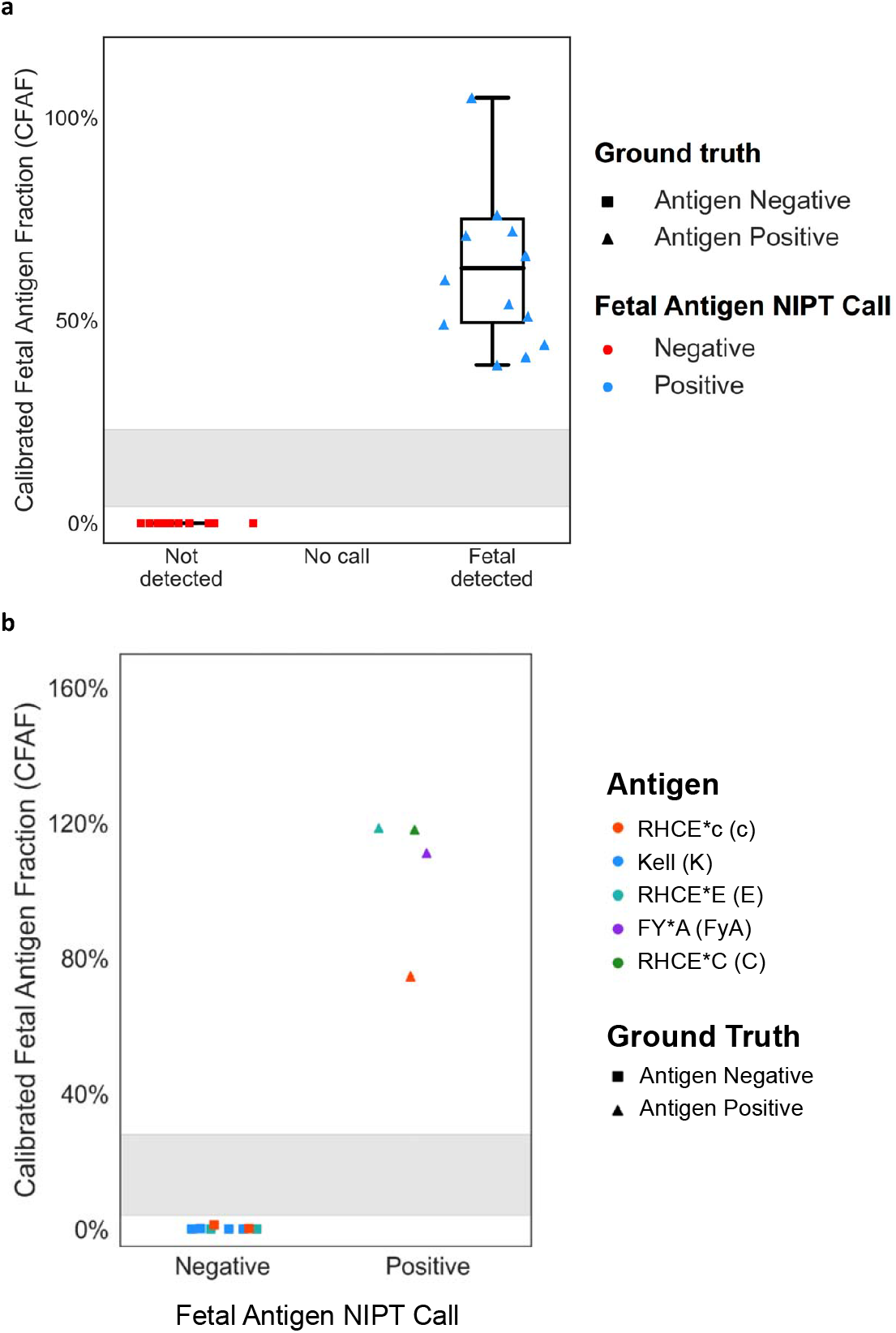
Calibrated fetal RhD is plotted against fetal fraction for 23 samples from the LIFECODES biobank with known fetal serology. All NIPT results were concordant with fetal serology. Antigen detected is indicated in blue, antigen not detected is indicated in red. Neonatal serology RhD-positive is indicated with a triangle and neonatal serology RhD-negative is indicated with a square.

The four samples from the UNITY fetal antigen patient registry included, (i) one participant with anti-c, (ii) one participant with anti-Kell and anti-Fy^a^ antibodies, (iii) one participant with anti-E antibodies, and (iv) one participant with anti-C antibodies. After delivery, neonatal samples were obtained and sent to a reference laboratory, Grifols, to determine the concordance between NIPT assay and the ground truth genotype results.

Both the Grifols antigen genotype assay and the NIPT assay can evaluate all fetal antigen genotypes for all antigens included on the panel, regardless of alloimmunization status (in clinical practice, only ordered antigen is reported). Therefore, concordance was assessed for 12 antigen evaluations across the four samples; including the five antigens for which the pregnant person was alloimmunized. The NIPT-predicted fetal antigen genotype and neonatal antigen genotype were concordant for all 12 evaluations. (**Figure 4b, Table S8b**).

## DISCUSSION

This study presents the development and validation of NIPT for fetal antigen detection appropriate for the diverse U.S. population with no partner sample required. NIPT for fetal antigen detection can decrease patient risk and anxiety, as well as healthcare burden, namely by reducing the administration of anti-RhD immunoglobulin, which is unnecessary for the up to 40% of RhD-negative pregnant individuals carrying an RhD-negative fetus. Moreover, for the approximately 1% of alloimmunized pregnant individuals, NIPT can prevent unnecessary fetal monitoring for the estimated 50-65% who are carrying an antigen-negative fetus [5].

Implementation of NIPT as standard of care for the U.S. pregnant population requires excellent detection to prevent sensitization in RhD-negative individuals and HDFN in alloimmunized pregnancies, high sensitivity for all ancestries to ensure feasibility and equity of care, and a cost-effective assay to limit healthcare burden [1, 27]. These features have been achieved with this NIPT assay which is completed in a cost-effective manner by running the assay in parallel with aneuploidy NIPT, a screen that is recommended for all pregnant individuals [28].

An important distinction of this assay from its European counterparts is its ability to determine fetal RhD status when non-*RHD* gene deletion variants are present in the cfDNA, which occur more frequently in non-European individuals [15, 16]. Most Europe-based NIPT assays use PCR technology for qualitative assessment of the presence or absence of select exons in the *RHD* gene and assume homozygous *RHD* gene deletion without assessment of fetal fraction [14]. While some of these assays have algorithms to determine likelihood a non-*RHD* gene deletion variant is present, they still cannot predict fetal Rh phenotype in the presence of a non-*RHD* gene deletion variant, which results in no-call rates as high as 15% [12, 19, 20, 26].

The combination of the QCT technology-based quantitative approach with NGS described in this work enables the assay to detect both the common *RHD* gene deletion and other *RHD* variants, including *RHDΨ* and *Rh-CE-D* hybrid genes, that result in an RhD-negative phenotype [22]. In the current study, a fetal RhD phenotype was predicted for all samples with a *RHDΨ* variant (3.6%) detected, illustrating the clinical utility of this assay in a diverse population. The assay had a sensitivity and specificity of 100% (95% CI: 99%-100%) for RhD and returned an informative result for 99.8% of samples when a single call threshold was applied, illustrating the efficiency of the assay to determine fetal Rh-phenotype.

The assay described here was designed with an absolute expected fetal antigen molecule (AEM) threshold to maximize sensitivity at very low fetal fractions. In the current study, the assay predicted fetal antigen genotype in pregnancies as early as 10 weeks’ gestation with fetal fractions as low as 1.1%. Current qualitative NIPT assays are unable to quantify fetal cfDNA to compare to an AEM threshold and therefore require a later gestational age; this can lead to unnecessary parental anxiety and pregnancy monitoring [14, 18]. The intermediate no-call CFAF threshold, where a fetal antigen status is not reported and a new sample is requested from the patient, maximizes the specificity of the assay. Even with the additional assurance steps, 99.5% of the NIPT assays on samples from pregnant individuals returned an informative result.

Fetal RhD detect by NIPT assay is challenging due to the many non-RHD gene deletions and RHD-CE-D hybrid genes that result in an RhD-negative phenotype and variants that result in grey-zone phenotypes. This limited assessment can result in referral of patients for NIPT who are not at risk for RhD sensitization [16]. Furthermore, the one-time serological evaluation that is standard of care for pregnant individuals does not evaluate RHD gene genotype and frequently is unable to detect the gray-zone phenotypes that can result in inadvertent, inappropriate referral of patients for RhD NIPT [1]. The current NIPT assay was designed to be robust to these edge cases. Specifically, the five amplicon based assay is designed to predict a fetal RhD-positive phenotype for rare RHD gene variants. This design ensures a low-risk misclassification (potentially unnecessary anti-RhD administration to individuals not at risk for RhD sensitization) for these edge cases.

The strengths of the assay are further illustrated in two biobank samples that did not meet the inclusion criteria. We still performed the NIPT analysis on them to assess the limits of the assay. The NIPT analysis of the sample from the suspected weak D pregnant individual predicted a fetal RhD-positive genotype. The neonatal serology showed RhD-negative. This discrepancy illustrates how the assay is designed to result in a low-risk misclassification that would recommend anti-RhD immunoglobulin administration in such weak D cases. For the purposes of understanding this case, the cfDNA sequencing data was interrogated and compound heterozygous *RHD* gene variants were identified (p.T201R and p.F223V) in the exon 4 amplified region [16]. This genotype is consistent with Weak D type 4, which explains the inconsistent pregnant individual serology. No wild type *RHD* was present in the cfDNA, suggesting the fetus and the pregnant parent had the same Weak D type 4 variants, indicating that neonatal serology might also result in inconsistent phenotypes. Similarly, the NIPT analysis of the second sample was excluded/no-called because it had only 0.15% fetal fraction. In this case, the NIPT analysis predicted a fetal RhD-positive result, consistent with the neonatal serology, demonstrating that the QCT design of the NIPT assay that quantifies RHD gene molecules can be robust to low fetal fractions.

This study is limited by a modest number of clinical samples with known neonatal antigen status and the late gestational age of some of these samples. Banked plasma from RhD-negative pregnancies with known neonatal Rh typing was readily available because Rh typing is routinely performed for all neonates born to RhD-negative individuals [1]. However, assessment of other RBC antigens is not routinely performed on neonates because it does not inform clinical care of the infant [29]. Therefore, this work includes the initial data from a prospective study to collect DNA samples for neonatal antigen genotyping.

## Conclusion

This novel NGS-based NIPT for the detection of fetal antigen genotypes demonstrated high performance. The NIPT results were 100% concordant with fetal antigen status when fetal or neonatal genotype or serology was known without the need for a partner sample. This assay addresses the ACOG-identified barriers of feasibility and inclusivity to implementing NIPT as the standard of care for RhD-negative individuals, allowing for selective use of antenatal anti-RhD immunoglobulin prophylaxis. Moreover, it would serve as an excellent tool for the targeted management of alloimmunized pregnancies at risk for HDFN [1].

## Supporting information

Sup Methods and Results

## Data Availability

All data produced in the present study are available upon reasonable request to the authors.

## Acknowledgments

We would like to thank Shan Riku, MBA, Oguzhan Atay, PhD, John ten Bosch, PhD, Kenneth J. Moise, MD, Cortney Cino, BA, Alexander Ni, PhD, Haley King, MSN, RN for their contributions. We would also like to thank the research participants.

## Author Contributions

BA and BPL made substantial contributions to the conception and design of the work, analysis, and interpretation of the data, drafted the manuscript, approved the submitted version and agree to be accountable for their contributions, and appropriately resolve any questions. SH, XB, AMB and DC made substantial contributions to the analysis, and interpretation of the data, revised the manuscript, approved the submitted version and agree to be accountable for their contributions, and appropriately resolve any questions. JC and BH made substantial contributions to the interpretation of the data, revised the manuscript, approved the submitted version and agree to be accountable for her contributions, and appropriately resolve any questions. JH made substantial contributions to the design of the study, revised the manuscript, approved the submitted version and agree to be accountable for their contributions, and appropriately resolve any questions. JW made substantial contributions to the interpretation of the data, to acquisition of samples, drafted the manuscript, approved the submitted version and agree to be accountable for their contributions, and appropriately resolve any questions. DEC and TFM made substation contributions to acquisition of samples, revised the manuscript, approved the submitted version and agree to be accountable for their contributions, and appropriately resolve any questions. KJG made substantial contributions to conception of the study and study design, acquisition of samples, revised the manuscript, approved the submitted version and agree to be accountable for her contributions, and appropriately resolve any questions.

## Data Availability

The minimal dataset that would be necessary to interpret, replicate and build upon the findings reported in the article is available upon request from the corresponding author.

## Ethics Declarations

Informed consent was obtained for all human research participants included in this publication.

