## Supplementary material for "Validation of a Non-invasive Prenatal Test for Fetal RhD, C, c, E, Kell and FyA Antigens": Sup Methods and Results

| **Table S1.** Calibrated fetal antigen fraction (CFAF) detected and not detected ranges and were determined through analysis of over 8K clinical samples. CFAFs that fall between Detected and Not Detected Ranges are reported out as a no call and in a clinical scenario a new sample is requested. Absolute expected fetal molecule (AEM) threshold was determined by Poisson distribution where at the lowest AEM the sensitivity does not fall below 98%. | | | | |
| --- | --- | --- | --- | --- |
| **Antigen** | **CFAF Antigen Not Detected Range** | **CFAF Antigen Detected Range** | **Fetal Fraction Automated Report Range**** | **AEM Threshold** |
| RhD | ≤4.0% | >23% | ≥1.5% | ≥2 |
| RhCE*C (C) | ≤5.0% | 24-300% | ≥1.5% | ≥6 |
| RHCE*c (c) | ≤7.0% | 26-300% | ≥1.5% | ≥4 |
| RHCE*E (E) | ≤5.0% | 24-300% | ≥1.5% | ≥6 |
| KEL*K (K) | ≤5.0% | 24-240% | ≥1.5% | ≥4 |
| FY*A (FyA) | ≤9.0% | 28-300% | ≥1.5% | ≥4 |
| Abbreviations: calibrated fetal antigen fraction (CFFAF), absolute expected fetal molecule (AEM) | | | | |

| **Table S2.** Coriell identities of the preclinical parent-child samples used for clinical validation. Samples were mixed to at various concentrations to mimic fetal fraction cfDNA samples ranting from 1.5-12%. | | | | | |
| --- | --- | --- | --- | --- | --- |
| **Sample Type** | **"Pregnant Person" Status** | **"Pregnant Person" Coriell ID** | **"Fetus" Status** | **"Fetus" Coriell ID** | **Total Number of Samples** |
| RhD Negative | RhD -/- | NA06989 | RhD -/- | NA12490 | 72 |
| RhD Negative | RhD -/- | NA12273 | RhD -/- | NA10837 | 72 |
| RhD Positive | RhD -/- | NA12156 | RhD +/- | NA10831 | 108 |
| RhD Positive | RhD -/- | NA12004 | RhD +/- | NA10838 | 108 |
| RhD Negative# | RhD -/- | HG01620 | RhD -/- | HG01621 | 48 |
| RhD Negative# | RhD -/- | NA19131 | RhD +/- | NA19132 | 48 |
| RhCE*c negative | RhCE*c -/- | HG00590 | RhCE*c -/- | HG00591 | 48 |
| RhCE*c positive | RhCE*c -/- | HG00566 | RhCE*c +/- | Hg00567 | 48 |
| RhCE*C negative | RhCE*C -/- | NA19131 | RhCE*C -/- | NA19132 | 48 |
| RhCE*C positive | RhCE*C -/- | HG00565 | RhCE*C +/- | HG00567 | 48 |
| RhCE*E negative | RhCE*E -/- | NA19656 | RhCE*E -/- | NA19654 | 48 |
| RhCE*E positive | RhCE*E -/- | HG00566 | RhCE*E +/- | HG00567 | 48 |
| KEL*K negative | K -/- | HG00565 | K -/- | HG00567 | 96 |
| KEL*K positive | K -/- | NA19656 | K +/- | NA19654 | 96 |
| FY*A negative | Fy^a^ -/- | HG01173 | Fy^a^ -/- | HG01175 | 48 |
| FY*A positive | Fy^a^ -/- | HG01522 | Fy^a^ +/- | HG01523 | 48 |

| **Table S3.** Characteristics of the 15,393 clinical samples where the pregnant person was negative for at least one antigen of interest. | | |
| --- | --- | --- |
|  | **Mean** | **Range** |
| Pregnant Individual's Age | 28.9 | 12-54 |
| Gestational Age (weeks) | 13.8 | 10-39 |
| Fetal Fraction | 8.8% | 1.5-37.8% |
| Race and Ethnicity* |  |  |
| White, Non-Hispanic | 4792 | 43.2% |
| Black, Non-Hispanic | 2051 | 18.5% |
| Hispanic | 2963 | 26.7% |
| Asian | 379 | 3.4% |
| Other | 905 | 8.2% |
| Unknown | 4303 |  |
| Total analyzed for each antigen# |  |  |
| RhD | 1673 | 10.9% |
| RhCE*C (C) | 5733 | 37.2% |
| RHCE*c (c) | 2981 | 19.4% |
| RHCE*E (E) | 10601 | 68.9% |
| KEL*K (K) | 14530 | 94.4% |
| FY*A (FyA) | 5937 | 38.6% |
| *Out of the 11,090 where race or ethnicity were known, N and % | | |
| #Only samples where the pregnant person was antigen negative by next generation sequencing were analyzed to mimic clinical use of the assay, N and % | | |

| **Table S4**. Proportion of pregnant people negative for each antigen based on NGS of cfDNA on 15,393 retained clinical samples. Samples are not mutually exclusive. Samples for each antigen did not pass the initial quality control due to fetal fraction or expected absolute molecular count were not analyzed. For the samples analyzed by fetal antigen NIPT, proportion of fetal antigen Detected, fetal Antigen Not Detected, or no call for fetal antigen. | | | | | |
| --- | --- | --- | --- | --- | --- |
| **Antigen** | **N** | **% Quality Control Fail** | **% NIPT Fetal Antigen Not Detected** | **% NIPT Fetal Antigen Detected** | **% Indeterminate Range No Call Fetal Antigen** |
| RhD | 1673 | 0.7% | 29.9% | 68.5% | 0.9% |
| RhCE*C (C) | 5733 | 5.1% | 60.9% | 33.4% | 0.6% |
| RHCE*c (c) | 2981 | 0.2% | 47.3% | 51.7% | 0.4% |
| RHCE*E (E) | 10601 | 3.1% | 80.3% | 16.1% | 0.6% |
| KEL*K (K) | 14530 | 1.6% | 95.7% | 2.5% | 0.2% |
| FY*A (FyA) | 5937 | 1.0% | 69.4% | 29.0% | 0.6% |
| Abbreviations: non-invasive prenatal testing (NIPT), cell free DNA (cfDNA), next generation sequencing (NGS) | | | | | |

| **Table S5.** Modeled mean, SD and sensitivity from truncated normal distribution fit to the NIPT CFAF values categorized as antigen detected from 15,939 plasma samples from pregnant individuals of unknown fetal antigen genotype and unknown pregnant person alloimmunization status who were negative (by genotype) for at least one of the RBC antigens. | | | |
| --- | --- | --- | --- |
| **Antigen** | **Modeled Mean** | **Modeled SD** | **Modeled Sensitivity** |
| RhD | 1.129 | 0.246 | 100% |
| RhCE*C (C) | 1.078 | 0.295 | 100% |
| RHCE*c (c) | 0.900 | 0.257 | 99.9% |
| RHCE*E (E) | 0.931 | 0.306 | 99.8% |
| KEL*K (K) | 1.034 | 0.367 | 99.6% |
| FY*A (FyA) | 0.995 | 0.324 | 99.8% |
| Abbreviations: Standard Deviation (SD) | | | |

| **Table S6**. 1,683 samples retained clinical samples with two samples. Samples are not mutually exclusive. Concordance of independent fetal antigen NIPT analysis run on each sample duo stratified by samples that were concordant for fetal antigen genotype positive and fetal antigen genotype negative. The direction of the one discordant sample is unknown as the true fetal antigen genotype is unknown for these samples. | | | | |
| --- | --- | --- | --- | --- |
| **Antigen** | **Concordant Positive N** | **Concordant Negative N** | **Discordant N** | **% Agreement** |
| RhD | 31 | 37 | 0 | 100.0% |
| RhCE*C (C) | 195 | 333 | 1 | 99.8% |
| RHCE*c (c) | 164 | 112 | 0 | 100.0% |
| RHCE*E (E) | 183 | 767 | 3 | 99.7% |
| KEL*K (K) | 29 | 1341 | 0 | 100.0% |
| FY*A (FyA) | 166 | 463 | 1 | 99.8% |

| **Table S7.** Characteristics of the 27 clinical samples with known neonatal antigen serology (RhD) or genotype | | |
| --- | --- | --- |
|  | **Mean** | **Range** |
| **Pregnant Individual's Age (yr)** | 35 | (23-44) |
| **Gestational Age (wk)** | 14.9 | (8.9-31.0) |
| **Fetal Fraction** | 7.5% | (1.2%-23.6%) |
| **Race and Ethnicity*** |  |  |
| Black, Non-Hispanic | 3 | 11.1% |
| Mixed, Non-Hispanic | 1 | 3.7% |
| Other, Hispanic | 4 | 14.8% |
| South Asian, Non-Hispanic | 1 | 3.7% |
| White, Non-Hispanic | 18 | 66.7% |

| **Table S8a.** Concordance of NIPT results and neonatal genotype or serology of the 4 UNITY study clinical samples from alloimmunized pregnant individuals and neonatal fetal antigen genotype and 23 biobank samples from RhD-negative pregnant individuals with neonatal serology. Detected (D), Not Detected (ND) | | | | | | | | | | |
| --- | --- | --- | --- | --- | --- | --- | --- | --- | --- | --- |
| **Antigen** | | **RhD** | **RhCE*C (C)** | | **RHCE*c (c)** | | **RHCE*E (E)** | **KEL*K (K)** | | **FY*A (FyA)** |
| **RhD^** | | 23:23 (12D, 11ND) |  | |  | |  |  | |  |
| **RhCE*C (C)** | |  | 1:1 (1D) | |  | |  |  | |  |
| **RHCE*c (c)** | |  |  | | 2:2 (1D, 1ND) | |  |  | |  |
| **RHCE*E (E)** | |  |  | |  | | 3:3 (2D, 1ND) |  | |  |
| **KEL*K (K)** | |  |  | |  | |  | 4:4 (4ND) | |  |
| **FY*A (FyA)** | |  |  | |  | |  |  | | 2:2 (2D) |
| For the alloimmunized participants, concordance for all antigens in which the pregnant individual was genotype negative (including the antigen the participant had antibodies to). | | | | | | | | | | |
| ^Biobank samples from RhD-negative pregnant individuals | | | | | | | | | | |
| **Table S8b.** Concordance of NIPT results and neonatal antigen genotype in the 4 UNITY study clinical samples from alloimmunized individuals (Figure 4b). | | | | | | | | | | |
| **Study ID** | **Pregnant person antibody** | | | **NIPT result** | | **Neonatal genotype result** | | |  |  |
| 1 | anti-C | | | C Detected,  K Not Detected | | C Detected,  K Not detected | | |  |  |
| 2 | anti-E | | | E Not Detected,  K Not Detected  c Not Detected, | | E Not Detected,  K Not Detected,  c Not Detected, | | |  |  |
| 3 | anti-K, anti-FyA | | | K Not Detected,  FyA Detected,  E Not Detected,  c Not Detected | | K Not Detected, FyA Detected,  E Not Detected,  c Not Detected | | |  |  |
| 4 | anti-c | | | c Detected,  E Detected,  K Not Detected | | c Detected,  E Detected,  K Not Detected | | |  |  |
